# Reduction in Spontaneous and Iatrogenic Preterm Births in Twin Pregnancies During COVID-19 Lockdown in Melbourne, Australia: A Multicenter Cohort Study

**DOI:** 10.1101/2023.05.16.23289144

**Authors:** Juliana M Manno, Melvin B Marzan, Daniel L Rolnik, Stephanie Potenza, Natasha Pritchard, Joanne M Said, Kirsten R Palmer, Clare L Whitehead, Penelope M Sheehan, Jolyon Ford, Ben W Mol, Susan P Walker, Lisa Hui

## Abstract

**Background:** Melbourne, Australia, recorded one of the longest and most stringent pandemic lockdowns in 2020, which was associated with an increase in preterm stillbirths among singleton pregnancies. Twin pregnancies may be particularly susceptible to the impacts of pandemic disruptions to maternity care due to their higher background risk of adverse perinatal outcomes.

**Objective:** To compare the rates of adverse perinatal outcomes in twin pregnancies exposed and unexposed to lockdown restrictions in Melbourne.

**Study Design:** Multicenter retrospective cohort study of all twin pregnancies birthing in public maternity hospitals in Melbourne. We compared perinatal outcomes between a pre-pandemic group (‘unexposed’) and two lockdown-exposed groups: exposure 1 from 22 March 2020 to 21 March 2021 and exposure 2 from 22 March 2021 to 27 March 2022. We analyzed routinely-collected maternity data on all twin births <u>≥</u>20 weeks where outcomes were available for both infants. The primary outcomes were rates of preterm birth<37 weeks and all-cause stillbirth. Multivariable log-binomial regression models were used to compare perinatal outcomes between the pre-pandemic group and women in whom weeks 20^+0^ to 40^+0^ of their pregnancy occurred entirely during each lockdown-exposure period. Perinatal outcomes were calculated per infant; maternal outcomes were calculated per pregnancy.

**Results:** We included 2267 women birthing twins. Total preterm births<37 weeks were significantly lower in the exposure 1 group compared with the pre-pandemic group (63.1% vs 68.3% respectively; adjusted risk ratio, aRR 0.92 95% CI 0.87-0.98, p=0.01). This was mainly driven by fewer spontaneous preterm births (18.9% vs 20.3%; aRR 0.95 95%CI 0.90- 0.99, p=0.04) and a trend to fewer iatrogenic preterm births (44.1% vs 48.1%; aRR 0.97 95%CI 0.92-1.03, p=0.39). There were also significantly lower rates of preterm birth<34 weeks in the exposure 1 group compared with the pre-pandemic group (19.9% vs 23.0%, aRR 0.93 95%CI 0.89-0.98 p=0.01). Total iatrogenic births for fetal compromise were significantly lower (13.4% vs 20.4%; aRR 0.94 95%CI 0.89–0.98, p=0.01). There were fewer special care nursery admissions (38.5% vs 43.4%; aRR 0.91 95%CI 0.87-0.95, p<0.001). There was no associated difference in all-cause stillbirths (1.5% vs 1.6%; aRR 1.00 95%CI 0.99-1.01, p=0.82), birthweight<3^rd^ centile (5.7% vs 6.0%; aRR 1.00, 95%CI 0.98-1.02 p=0.74) or neonatal intensive care unit admissions in the exposure 1 group compared to the pre-pandemic group. In contrast, when comparing the pre-pandemic group with exposure 2 group, there was no significant difference in the rates of preterm birth<37 or <34 weeks. However, during exposure 2 the rate of preterm birth<28 weeks was significantly higher (7.2% vs 4.8%; aRR 1.03 95%CI 1.01-1.05, p=0.04) and infants were more likely to be admitted to a neonatal intensive care unit (25.0% vs 19.6%; aRR 1.06 95%CI 1.03-1.10, p<0.0001) compared with the pre-pandemic period.

**Conclusions:** Melbourne’s first lockdown-exposure period was associated with fewer twin preterm births<34 and <37 weeks without significant differences in stillbirths or adverse newborn outcomes. These lower rates were not sustained in the second exposure period. Pandemic conditions may provide important lessons for future antenatal care of twin pregnancies, including prevention of preterm birth and optimal timing of birth.

## INTRODUCTION

Melbourne, Australia experienced one of the longest cumulative COVID-19 lockdowns over 2020 and 2021.^1^ The so-called ‘zero-COVID’ public health approach in 2020 successfully suppressed COVID-19 case numbers,^2^ creating an opportunity to examine the indirect impacts of lockdown on perinatal outcomes uncoupled from the effects of COVID-19 infections.

During the most stringent restrictions in 2020, mask mandates and a daily curfew were introduced, and residents were only permitted to leave home for four reasons. These included shopping for essential goods, one-hour-a-day of exercise, authorized work and medical care.^3^ Modifications to antenatal care included the introduction of telehealth, increasing the interval between face-to-face obstetric visits and the rationalization of ultrasound appointments.^4^ During 2021, shorter lockdowns were intermittently reintroduced to control outbreaks, in concert with a national vaccine program commencing in March 2021.^5^ There was a gradual loosening of restrictions outlined in a ‘roadmap’ based on reaching vaccination coverage targets.^6^

We previously reported that 2020 lockdown conditions were associated with reduced iatrogenic preterm birth (PTB) for fetal compromise in singletons and increased preterm stillbirths.^7^ Twin pregnancies may be more vulnerable than singletons to disruptions in maternity care due to their higher risk of adverse outcomes.^8, 9^ Stillbirths are increased thirteen-fold in monochorionic and five-fold in dichorionic twin pregnancies compared to singletons.^10^ Consequently, current guidelines recommend elective delivery before 37^+6^ weeks for dichorionic twins, 36^+6^ weeks for monochorionic diamniotic (MCDA) twins, and 32^+0^-33^+6^ weeks for monochorionic monoamniotic (MCMA) twins to prevent unexpected antepartum stillbirths.^9, 11^ Similarly, more frequent ultrasounds are recommended due to the increased risk of fetal growth restriction (FGR) and other complications including twin-twin transfusion syndrome (TTTS) in monochorionic twins.^12^

Most studies examining perinatal outcomes during the pandemic excluded multiple pregnancies or combined them with singletons rendering a gap in the literature on the impact of COVID-19 lockdowns on perinatal outcomes in twins. The Collaborative Maternity and Newborn Dashboard (CoMaND) for the COVID-19 pandemic was established in Melbourne to monitor the impact of lockdown on mothers and infants. We used CoMaND project data to investigate the impact of Melbourne’s lockdown on the rates of PTB, stillbirth and other maternal and perinatal outcomes in twin pregnancies.

## MATERIALS AND METHODS

### Setting

This multicenter retrospective cohort study obtained routinely-collected data on all twin births in public maternity hospitals in metropolitan Melbourne from 1 January 2018 to 27 March 2022. The twelve participating hospitals included all four tertiary centers caring for high-risk and extremely premature pregnancies.

The study period was divided into a pre-pandemic (weeks commencing 1 January 2018 to 8 March 2020) and two pandemic-exposure periods. Exposure 1 was defined as the period where the National Stringency Index was continuously ≥50 on the Oxford COVID-19 Government Response Tracker scale,^13^ in keeping with international definitions for significant lockdown restrictions.^14^ This corresponded to the weeks commencing 22 March 2020 to 21 March 2021. Exposure 2 was defined as the period following exposure 1 to the week commencing 27 March 2022. We divided the exposure period into two epochs as they were characterized by a shift from the ‘COVID-zero’ strategy to the less stringent ‘living-with-COVID’ approach.

### Data Sources and collection

We extracted de-identified patient-level data from the electronic birthing outcomes system (BOS, version 6.04) or equivalent at each hospital.^15^ Only births≥20^+0^ weeks are routinely collected. For privacy protection, hospital data managers converted the infant date-of-birth into the ordinal calendar week-of-birth (1-52). The first day-of-the week was designated Sunday.

Given minor variations in data collection between hospitals, correct mother-twin pairs were manually matched using a birth episode code. Where this code was missing, we performed manual probabilistic pairing of births using week-of-birth and maternal characteristics (country-of-birth, age, height, postcode, obstetric complications). Unpaired infants were excluded. As chorionicity information was incomplete, we planned a subgroup analysis by sex-concordance (sex-discordant twins presumed dichorionic; sex-concordant twins presumed either dichorionic or monochorionic).

### Inclusion and exclusion criteria

We excluded infants with birthweight<150g and births to non-Victorian residents. Births in private maternity hospitals (approximately 20% of births in 2020) and planned homebirths (<1% of births) were unavailable.^16^

We included infants with congenital anomalies or complications associated with monochorionicity (e.g. TTTS) as these are important contributors to the excess perinatal losses in twin pregnancies. Similarly, terminations of pregnancy (TOP) were not excluded as these may have been impacted by delayed diagnosis of fetal anomalies, reduced access to TOP<20^+0^ weeks or fetal therapy, or increases in maternal psychosocial stressors during the pandemic. We therefore use the term ‘all-cause stillbirths’ in our total cohort.

We performed a sensitivity analysis excluding births<24 weeks, TOP, and infants with congenital anomalies to generate a stillbirth rate for those pregnancies where iatrogenic preterm birth for fetal indications could be reasonably expected.

### Cohort definitions

In keeping with our previous work,^7^ we used the calculated week-of-last menstrual period (cLMP), rather than week-of-birth, to define the exposed cohort to avoid the fixed-cohort bias.^17^ The limitation of using date-of-birth to define a pregnancy cohort is that it fails to capture women who conceived at a similar time to other women in the cohort (and therefore had comparable lockdown exposure) but gave birth before the starting date. Similarly, lockdown-exposed pregnancies birthing after the study end date are missed, leading to potentially biased estimates of the exposure effect. We defined the exposed cohort to include women whose exposure to lockdown commenced no later than 20^+0^ weeks and who birthed at any gestation up to 40^+0^ weeks by the end of the data collection period We back-calculated a woman’s week-of-LMP using the sham week-of-birth and gestational age (GA) in completed weeks at delivery. The formula used for the cLMP was *first day of week-of-LMP=(week-of-birth–(GA×7))*. GA is expressed as weeks post-LMP, not post-conception. For example, for a woman birthing in the calendar week commencing Sunday 11 March 2018 at 37^+0^ weeks, her cLMP would be 25 June 2017. This woman would be 30^+1^ weeks at study commencement on 1 Jan 2018 and would therefore be excluded from the pre-pandemic cohort to avoid the aforementioned fixed-cohort bias.

We defined the pre-pandemic group as women with cLMP from 13 August 2017 to 16 June 2019. This includes women for whom weeks 20^+0^ to 40^+0^ occurred in the pre-pandemic period (Figure 2). The first exposed group comprised women for whom weeks 20^+0^ to 40^+0^ occurred during exposure 1 (cLMP from weeks commencing 3 November 2019 to 14 June 2020, inclusive). The exposure 2 group followed directly on from exposure 1 (cLMP from 21 June 2020), and concluded with women with cLMP in the week of 20 June 2021, ensuring a minimum of 40^+0^ weeks had elapsed by the end of our data-collection period on 31 March 2022 (Figure 2). The exposure 2 group did not exclude women whose pregnancies overlapped with the exposure 1 period.

**Figure 1:**
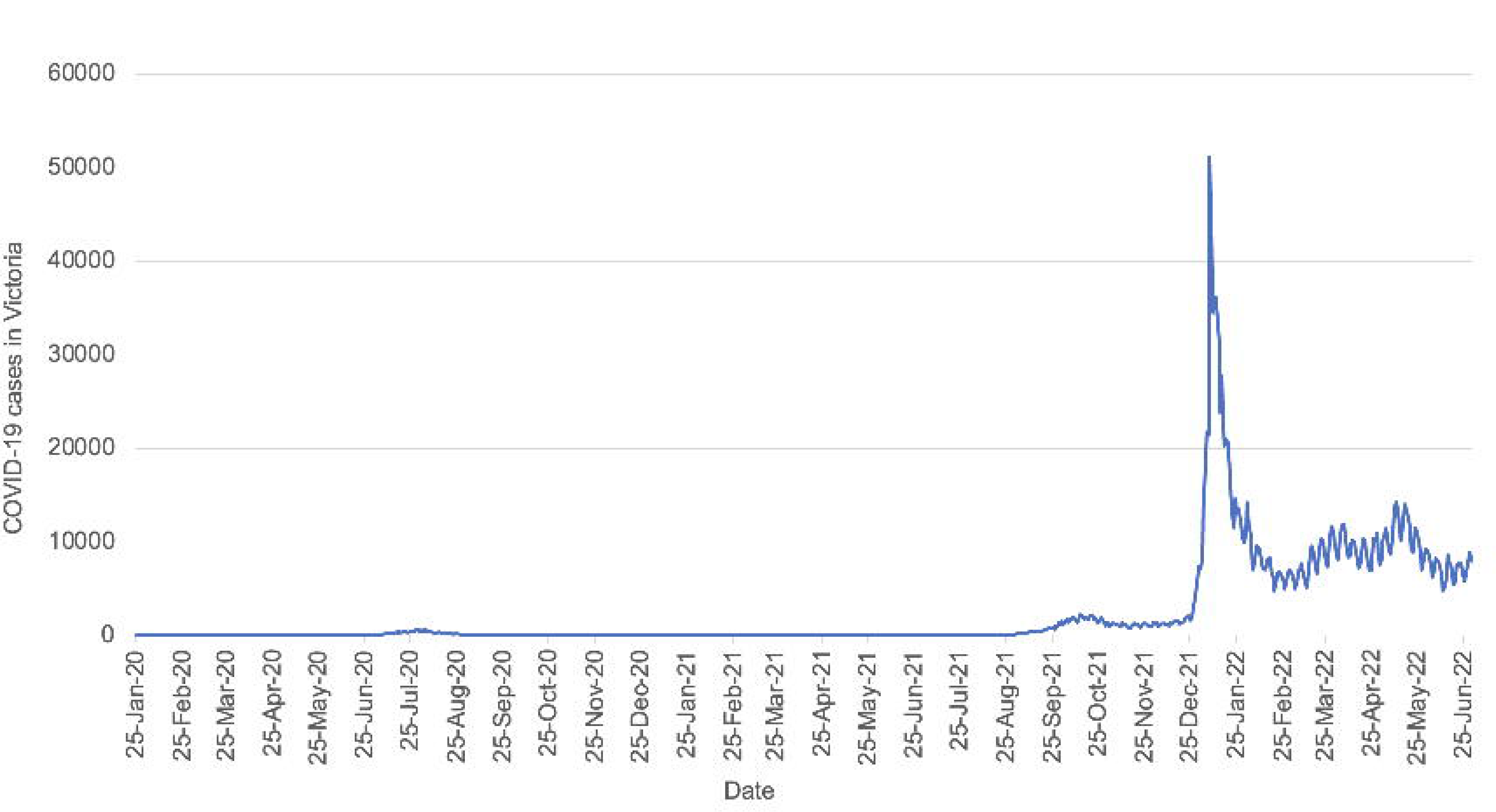
COVID-19 case numbers in Victoria. Data source: https://www.coronavirus.vic.gov.au/victorian-coronavirus-covid-19-data. Accessed on 7 September 2022.

**Figure 2:**
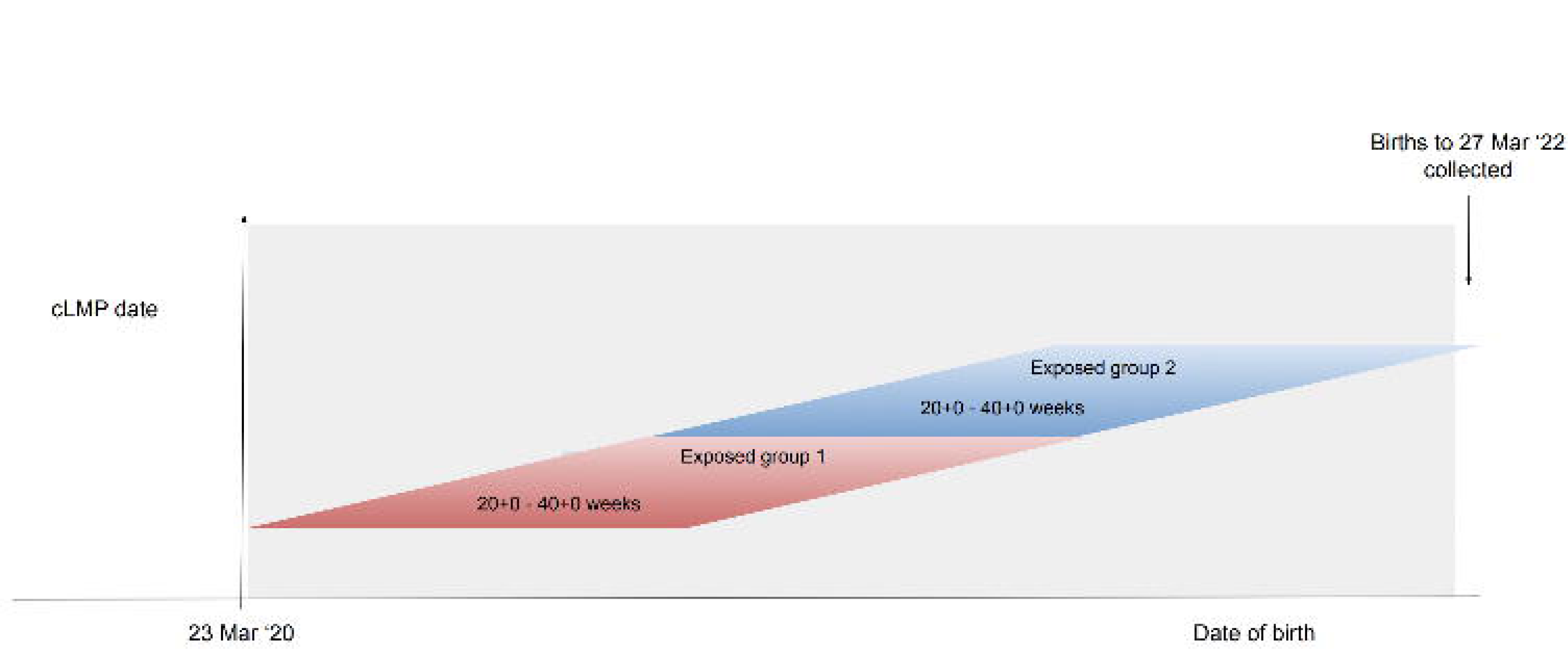
Pandemic-exposed groups 1 and 2 timeline.

### Outcomes

#### Primary outcomes

1. PTB<37 weeks; total, iatrogenic and spontaneous (denominator: number of pregnancies). Iatrogenic birth was defined as a birth following induction of labor (IOL) or birth by cesarean section (CS) with no labor.
2. 2. All-cause stillbirths (denominator: number of infants)

#### Secondary outcomes (denominator: number of pregnancies)

1. Total, iatrogenic and spontaneous

a. PTB<34 weeks
b. PTB<28 weeks
c. PTB<24 weeks
2. Iatrogenic birth for suspected fetal compromise; total, term≥37 weeks, and preterm<37 weeks. This was defined as any iatrogenic birth performed for suspected FGR, oligohydramnios, abnormal umbilical artery Doppler studies, placental insufficiency, abnormal cardiotocography, fetal distress with no labor, or reduced fetal movements^18^
3. Mode of birth; vaginal, CS after labor onset, CS with no labor
4. First antenatal visit≤12 weeks, defined as the first planned antepartum visit to a midwife or doctor (community or hospital)
5. Severe post-partum hemorrhage (estimated blood los ≥1000ml)

#### Secondary outcomes (denominator: number of infants)

i. 6. FGR, defined as birthweight≤3^rd^ centile using Australian population-based sex-specific birthweight charts^19^
ii. 7. Special care nursery (SCN) admission
iii. 8. Neonatal intensive care unit (NICU) admission
iv. 9. 5-minute Apgar scores<7 (term infants only)
v. 10. Congenital anomalies, excluding minor anomalies in accordance with state government practice^20^
vi. 11. Stillbirth rate (excluding births <24 weeks, TOPs and congenital anomalies)

#### Subgroup analysis

Sex-discordant twins were compared with sex-concordant twins for these selected outcomes: median GA, birthweight, all-cause stillbirth, PTB<37 weeks and iatrogenic birth for suspected fetal compromise.

### Statistical Analyses

Data were analyzed in STATA SE v17.^21^ Maternal demographics were tested with one-way analysis of variance (ANOVA) for continuous variables and chi-squared tests of independence for categorical variables. An interaction term between pregnancy smoking status and cohort categories was added because smoking may be in the causal pathway between exposure and the adverse outcomes studied. Socioeconomic status was derived from the maternal postcode and assigned an index of relative socioeconomic advantage and disadvantage (IRSAD) quintile using the Australian Bureau of Statistics (ABS) score.^22^ Region of birth and interpreter requirement were used as proxy indicators of ethnicity given the limitations of self-reported ethnicity in our population.^23^ Maternal COVID-19 infection at any time during pregnancy was extracted by text mining of data entered under maternal medical conditions, obstetric complications and labor complications. Statistical significance was set at p<0.05. Maternal COVID-19 vaccination coverage was only recorded during exposure 2 as the Australian vaccine roll-out commenced in March 2021, with active recommendations for antenatal vaccination announced on 9 June 2021. However, many pregnant women were ineligible based on their age and the absence of other medical conditions.^24^ Mandatory data-collection on maternal COVID-19 vaccination status commenced from 1 July 2021 and pregnant women were prioritized in the rollout from 22 July 2021.^25^

Outcomes in the pre-pandemic cohort were independently compared with those of exposures 1 and 2. Cohort comparisons were made with multivariable log-binomial regression models adjusted for covariates of PTB including maternal age, first measured body mass index, region of birth (ABS classifications), interpreter requirement, parity, socioeconomic status, smoking status, and pertussis vaccination. Effect estimates are presented as adjusted risk ratios (aRR) with 95% confidence intervals (95%CI).

Missing covariates data were accounted by implementing the multiple imputation by chained equations using the “mi impute” command in STATA.^26^

### Sensitivity analysis

We performed two sensitivity analyses to better understand the influences of the exposures on the primary outcomes.

i. As TOPs and congenital anomalies may be on the causal pathways between exposure and our primary outcomes, we performed descriptive and regression analysis for the primary outcome measures (PTB<37 weeks, and stillbirths) excluding births<24 weeks, pregnancies with one or more infants with a congenital anomaly, TOP or selective feticide.
ii. We also performed a sensitivity analysis on the total cohort (including births<24 weeks, TOPs, and infants with congenital anomalies) after excluding sex-

discordant twins to examine the robustness of our results in a group enriched for monochorionic twins.

### Ethics

Ethics approval was obtained from Austin Health (HREC/64722/Austin-2020) and Mercy Health (ref. 2020-031). As a retrospective study of routinely-collected non-identifiable data, a consent waiver was granted.

## RESULTS

We included 2267 women birthing twins: 1219 in the pre-pandemic cohort and 1048 in the pandemic-exposed cohort: 433 in exposure 1 and 615 in exposure 2 (Figure 3). Maternal characteristics are shown in Table 1. There was a significant difference in smoking during pregnancy, being significantly higher in exposure 1 compared to the pre-pandemic and exposure 2 groups (10.1% vs 6.3% vs 5.8%, respectively, p=0.04). Maternal pertussis and influenza vaccinations were lower in each exposure group compared with the pre-pandemic group (72.1% vs 85.6%, p<0.001, and 70.2% vs 75.9%, p<0.001, respectively). Five women (0.8%) were infected with SARS-CoV-2 in exposure 2 group, 2 women<37 weeks, 1 woman<34 weeks and 2 women<32 weeks, compared with no infections in the exposure 1 group (p=0.001). Only women in the exposure 2 group received a COVID-19 vaccination during pregnancy (55.0%).

**Figure 3:**
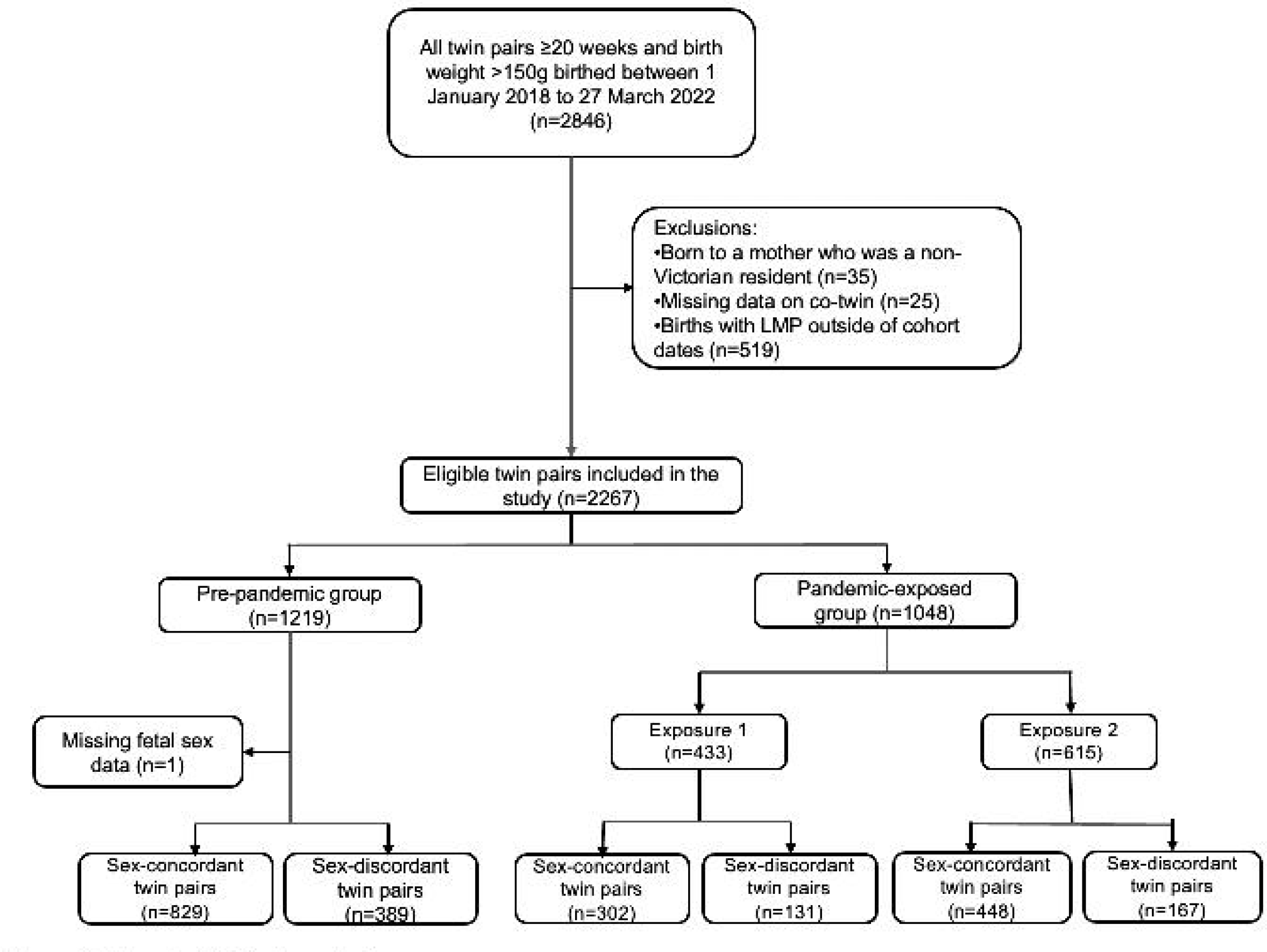
Flowchart of study population.

**Figure 4:**
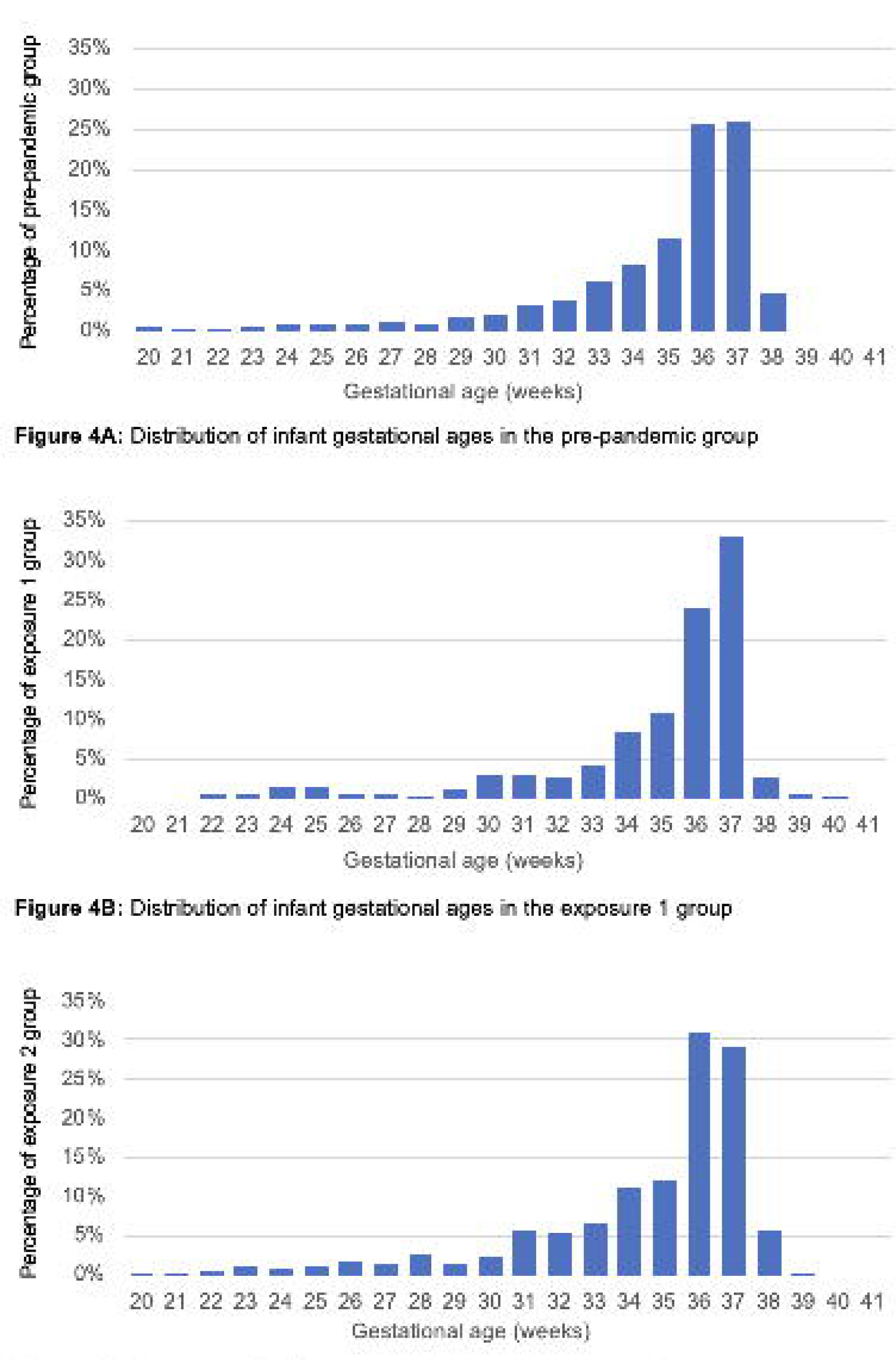
Distribution of infant gestational ages in the (A) Pre-pandemic, (B) Exposure 1 and (C) Exposure 2 groups, per pregnancy

**Table 1:** Comparison of maternal and pregnancy characteristics among the pre-pandemic, exposure 1 and exposure 2 groups

| Maternal characteristics | Pre-pandemic | Exposure 1 | Exposure 2 | P value* |
| --- | --- | --- | --- | --- |
|  | n = 1219 | n=433 | n=615 |  |
| Maternal age in years, mean (SD) | 32.72 (5.2) | 32.45 (4.6) | 32.27 (4.9) | 0.18 |
| Weight in kg, mean (SD) | 72.04 (21.2) | 69.78 (23.8) | 70.84 (22.6) | 0.17 |
| Height in cm, mean (SD) | 160.80 (24.1) | 157.23 (33.0) | 158.78 (30.3) | 0.05 |
| Smoking in pregnancy, n (%) | 58 (6.3) | 33 (10.1) | 26 (5.8) | 0.04 |
| Interpreter required, n (%) | 47 (3.9) | 15 (3.5) | 22 (3.6) | 0.92 |
| Pertussis vaccination during pregnancy, n (%) | 997 (85.6) | 308 (72.1) | 450 (74.5) | <0.001 |
| Influenza vaccination during pregnancy, n (%) | 887 (75.9) | 302 (70.2) | 367 (61.0) | <0.001 |
| ≥1 COVID-19 vaccine dose before or during pregnancy, n/N (%) | - | - | 197/358<sup>A</sup> (55.0) | NA |
| COVID-19 infection during pregnancy | - | 0 (0.0) | 5 (0.8) | 0.001 |
| <37 wk | - | 0 (0.0) | 2 (40.0) |  |
| <34 wk | - | 0 (0.0) | 1 (20.0) |  |
| <32 wk | - | 0 (0.0) | 2 (40.0) |  |
| <28 wk | - | 0 (0.0) | 0 (0.0) |  |
| Maternal age group in years, n (% of total) |  |  |  |  |
| 18-24 | 107 (8.8) | 40 (9.2) | 69 (11.3) | 0.33 |
| 25-29 | 247 (20.3) | 84 (19.4) | 119 (19.4) |  |
| 30-34 | 490 (40.2) | 185 (42.7) | 247 (40.4) |  |
| 35-39 | 280 (23.0) | 103 (23.8) | 143 (23.4) |  |
| ≥ 40 | 95 (7.8) | 21 (4.9) | 34 (5.6) |  |
| BMI categories in Kg/m<sup>2</sup>, n (% of total) |  |  |  |  |
| <18 | 13 (1.1) | 2 (0.5) | 3 (0.6) | 0.67 |
| 18 – 24 | 476 (40.6) | 169 (42.4) | 234 (44.0) |  |
| 25 – 29 | 370 (31.5) | 115 (28.8) | 146 (27.4) |  |
| 30 – 34 | 176 (15.0) | 63 (15.8) | 92 (17.3) |  |
| 35 – 39 | 83 (7.1) | 32 (8.0) | 35 (6.6) |  |
| ≥ 40 | 56 (4.8) | 18 (4.5) | 22 (4.1) |  |

| Region of birth, n (% of total pregnancies) |  |  |  |  |
| --- | --- | --- | --- | --- |
| Americas | 17 (1.4) | 9 (2.1) | 19 (3.1) |  |
| Australia and Associated Territories | 728 (59.8) | 254 (59.4) | 354 (58.0) |  |
| North Africa and Middle East | 64 (5.3) | 15 (3.5) | 28 (4.6) |  |
| North East Asia | 35 (2.9) | 12 (2.8) | 16 (2.6) |  |
| North West Europe | 38 (3.1) | 21 (4.9) | 18 (3.0) |  |
| Oceania | 46 (3.8) | 15 (3.5) | 19 (3.1) |  |
| South East Asia | 59 (4.8) | 22 (5.1) | 28 (4.6) |  |
| Southern and Central Asia | 167 (13.7) | 60 (14.0) | 96 (15.9) | 0.52 |
| Southern and Eastern Europe | 18 (1.5) | 6 (1.4) | 14 (2.3) |  |
| Sub-Saharan Africa | 46 (3.8) | 14 (3.3) | 17 (2.8) |  |
| Parity, n (%) |  |  |  |  |
| 0 | 569 (46.8) | 197 (45.5) | 293 (47.6) |  |
| ≥ 1 | 648 (53.3) | 236 (54.5) | 322 (52.4) | 0.79 |
| SEIFA quintile, n (%) |  |  |  |  |
| 1 – Most disadvantaged | 240 (19.7) | 89 (20.6) | 137 (22.3) |  |
| 2 | 207 (17.0) | 58 (13.4) | 83 (13.5) |  |
| **3** | 306 (25.1) | 101 (23.3) | 153 (24.9) | 0.18 |
| **4** | 279 (22.9) | 97 (22.4) | 140 (22.8) |  |
| **5 – Most advantaged** | 185 (15.2) | 88 (20.3) | 102 (16.6) |  |
| **Pregnancy characteristics, n (%)** |  |  |  |  |
| **Sex-concordant** | 829 (68.0) | 302 (69.8) | 448 (72.9) |  |
| **Sex-discordant** | 389 (32.0) | 131 (30.3) | 167 (27.2) | 0.11 |

*BMI*, body mass index; *SD*, standard deviation; *SEIFA*, socioeconomic index for areas.
Missing or implausible for weight=85 (3.76%); missing or implausible height=15 (0.66%), smoking status=0 (0%), need for interpreter variable=0 (0%), maternal age=6 (0.13%), BMI=169 (7.34%), region of birth=11 (0.49%), SEIFA=35 (1.55%), sex=1 (0.04%).
<sup>A</sup>The denominator for COVID-19 vaccination status is lower than total women in Exposure 2 group as vaccination status was only available from 1 July 2021.

Twin pregnancy outcomes are summarized in Table 2 and neonatal outcomes in Table 3.

**Table 2:** Primary and secondary outcomes for all pregnancies in the pre-pandemic, exposure 1 and exposure 2 cohorts

| Outcomes | Pre-pandemic<br>n = 1219 |  | Exposure 1<br>n=433 |  | Exposure 2<br>n=615 |  | Exposure 1 aRR |  |  | Exposure 2 aRR |  |  |
| --- | --- | --- | --- | --- | --- | --- | --- | --- | --- | --- | --- | --- |
|  | n | % | n | % | n | 27% | aRR | 95% CI | P value | aRR | 95% CI | P value |
| **Preterm birth <37 wk** |  |  |  |  |  |  |  |  |  |  |  |  |
| Total | 833 | 68.3 | 273 | 63.1 | 438 | 71.2 | 0.92 | 0.87-0.98 | 0.01 | 1.03 | 0.98-1.09 | 0.19 |
| Spontaneous | 247 | 20.3 | 82 | 18.9 | 136 | 22.1 | 0.95 | 0.90-0.99 | 0.04 | 1.00 | 0.95-1.04 | 0.83 |
| Iatrogenic | 586 | 48.1 | 191 | 44.1 | 302 | 49.1 | 0.97 | 0.92-1.03 | 0.39 | 1.04 | 0.99-1.10 | 0.16 |
| **Preterm birth <34 wk** |  |  |  |  |  |  |  |  |  |  |  |  |
| Total | 280 | 23.0 | 86 | 19.9 | 168 | 27.3 | 0.93 | 0.89-0.98 | 0.01 | 1.04 | 0.99-1.09 | 0.09 |
| Spontaneous | 129 | 10.6 | 45 | 10.4 | 72 | 11.7 | 0.97 | 0.94-1.01 | 0.15 | 1.01 | 0.98-1.04 | 0.65 |
| Iatrogenic | 151 | 12.4 | 41 | 9.5 | 96 | 15.6 | 0.96 | 0.92-1.00 | 0.05 | 1.03 | 1.00-1.07 | 0.09 |
| **Preterm birth <28 wk** |  |  |  |  |  |  |  |  |  |  |  |  |
| Total | 58 | 4.8 | 23 | 5.3 | 44 | 7.2 | 1.00 | 0.97-1.03 | 0.95 | 1.03 | 1.01-1.05 | 0.04 |
| Spontaneous | 30 | 2.5 | 12 | 2.8 | 22 | 3.6 | 1.00 | 0.98-1.02 | 0.79 | 1.02 | 1.00-1.03 | 0.09 |
| Iatrogenic | 28 | 2.3 | 11 | 2.5 | 22 | 3.6 | 0.98 | 0.96-1.00 | 0.08 | 1.00 | 0.98-1.02 | 0.94 |
| **Preterm birth <24 wk** |  |  |  |  |  |  |  |  |  |  |  |  |
| Total | 20 | 1.6 | 7 | 1.6 | 15 | 2.4 | 0.99 | 0.97-1.00 | 0.10 | 1.00 | 0.98-1.01 | 0.64 |
| Iatrogenic | 12 | 1.0 | 6 | 1.4 | 8 | 1.3 | 1.00 | 0.98-1.01 | 0.44 | 1.00 | 0.99-1.01 | 0.47 |
| Spontaneous | 8 | 0.7 | 1 | 0.2 | 7 | 0.8 | 0.99 | 0.98-1.00 | 0.10 | 1.00 | 0.99-1.01 | 0.88 |
| **Iatrogenic birth for suspected fetal compromise** |  |  |  |  |  |  |  |  |  |  |  |  |
| Total | 249 | 0.4 | 58 | 13.4 | 122 | 19.8 | 0.94 | 0.89-0.98 | 0.01 | 1.00 | 0.96-1.04 | 0.92 |
| ≥37 wk | 53 | 13.7 | 11 | 6.9 | 27 | 15.3 | 0.94 | 0.88-1.01 | 0.07 | 1.02 | 0.96-1.09 | 0.54 |
| <37 wk | 196 | 23.5 | 47 | 17.2 | 95 | 21.7 | 0.94 | 0.89-1.01 | 0.08 | 0.99 | 0.94-1.04 | 0.61 |
| **Mode of birth** |  |  |  |  |  |  |  |  |  |  |  |  |
| Vaginal | 361 | 29.6 | 112 | 25.9 | 180 | 29.3 | 0.93 | 0.88-0.98 | 0.01 | 0.96 | 0.92-1.01 | 0.12 |
| Cesarean section after<br>labor onset | 413 | 33.9 | 145 | 33.5 | 235 | 38.2 | 0.97 | 0.92-1.03 | 0.30 | 1.03 | 0.98-1.08 | 0.32 |
| Cesarean section with<br>no labor | 445 | 36.5 | 176 | 40.6 | 200 | 32.5 | 1.11 | 1.05-1.18 | 0.001 | 1.01 | 0.96-1.06 | 0.66 |
| **Pregnancy care indicators** |  |  |  |  |  |  |  |  |  |  |  |  |
| First antenatal visit<br>≤12 wk | 717 | 58.8 | 294 | 67.9 | 426 | 69.3 | 1.06 | 1.00-1.12 | 0.05 | 1.06 | 1.01-1.12 | 0.02 |
| Severe PPH > 1000ml | 129 | 10.6 | 40 | 9.2 | 66 | 10.7 | 0.96 | 0.92-0.99 | 0.02 | 0.98 | 0.95-1.01 | 0.23 |

**Table 3:** Infant outcomes in the pre-pandemic, exposure 1 and exposure 2 cohorts

| Outcomes | Pre-pandemic |  | Exposure 1 |  | Exposure 2 |  | Exposure 1 aRR |  |  | Exposure 2 aRR |  |  |
| --- | --- | --- | --- | --- | --- | --- | --- | --- | --- | --- | --- | --- |
|  | n=2438 | 58% | n=866 | 19% | n=1230 | 27% | aRR | 95%CI | P value | aRR | 95%CI | P value |
| All-cause stillbirths | 38 | 1.6 | 13 | 1.5 | 26 | 2.1 | 1.00 | 0.99-1.01 | 0.82 | 1.00 | 0.99-1.01 | 0.63 |
| Fetal growth restriction | 145 | 6.0 | 49 | 5.7 | 70 | 5.7 | 1.00 | 0.98-1.02 | 0.74 | 1.00 | 0.98-1.02 | 0.88 |
| SCN admissions | 1058 | 43.4 | 333 | 38.5 | 431 | 35.0 | 0.91 | 0.87-0.95 | <0.001 | 0.87 | 0.84-0.91 | <0.001 |
| NICU admissions | 478 | 19.6 | 171 | 19.8 | 307 | 25.0 | 0.99 | 0.95-1.02 | 0.45 | 1.06 | 1.03-1.10 | <0.001 |
| 5-minute Apgar <7 | 28 | 3.6 | 16 | 5.0 | 20 | 5.7 | 1.00 | 0.97-1.03 | 0.91 | 1.01 | 0.98-1.04 | 0.38 |
| (term infants) |  |  |  |  |  |  |  |  |  |  |  |  |
| Congenital anomalies* | 106 | 4.4 | 40 | 4.6 | 49 | 4.0 | 0.98 | 0.97-1.00 | 0.06 | 0.97 | 0.96-0.99 | <0.001 |

*aRR*, adjusted risk ratio; *CI*, confidence interval; *SCN*, special care nursery; *NICU*, neonatal intensive care unit.

The outcomes of sex-concordant twins are compared with sex-discordant twins in Table 4.

**Table 4:** Comparison of birth outcomes between the sex-discordant and sex-concordant twins (combined pre-pandemic and exposure 1 and 2 cohorts)

| Per infant* | Infants from sex concordant pairs | Infants from sex discordant pairs | P value |
| --- | --- | --- | --- |
|  | n=3158 | n=1375 |  |
| Median gestational age in wk, days<sup>a</sup> | 36<sup>+0</sup> | 36<sup>+6</sup> | <0.0001 |
| Birthweight in g, mean (SD) | 2202.7 (673.6) | 2352.0 (640.7) | <0.0001 |
| All-cause stillbirths, n (%) | 60 (1.9) | 17 (1.2) | 0.11 |
| Preterm birth 37 wk, n (%) | 2309 (73.1) | 779 (56.7) | <0.001 |
| Iatrogenic birth for suspected fetal compromise, n (%) | 613 (19.4) | 217 (15.8) | <0.001 |

*SD*, standard deviation; wk, weeks.
<sup>a</sup>The rank sum test was used to calculate the P value for the median gestational age of twins.
\*1 pair with missing information on sex concordance are excluded. The denominator used was total infants (livebirths + stillbirths)

### Exposure 1 group versus pre-pandemic group

#### Primary outcomes

The rate of PTB<37 weeks was significantly lower in exposure 1 group compared with the pre-pandemic group (63.1% vs 68.3%; aRR 0.92, 95%CI 0.87-0.98, p=0.01) (Table 2). This was driven by fewer spontaneous PTBs (18.9% vs 20.3%; aRR 0.95, 95%CI 0.90-0.99, p=0.04) and a trend to fewer iatrogenic PTBs (44.1% vs 48.1%; aRR 0.97, 95%CI 0.92-1.03, p=0.39) (Table 3).

All-cause stillbirth rates were not significantly different between the groups (1.5% vs 1.6%, aRR 1.00, 95%CI 0.99-1.00, p=0.82) (Table 3).

#### Secondary outcomes

There were significantly lower rates of PTB<34 weeks during exposure 1 compared with the pre-pandemic period (19.9% vs 23.0%, aRR 0.93, 95%CI 0.89-0.98, p=0.01) mediated by a trend towards fewer iatrogenic births<34 weeks. Total iatrogenic births for the specific indication of suspected fetal compromise were significantly lower (13.4% vs 20.4%; aRR 0.94, 95%CI 0.89–0.98, p=0.01) (Table 2).

The exposure 1 group had more CS with no labor (40.6% vs 36.5%; aRR 1.11, 95%CI 1.05-1.18, p=0.001) and correspondingly fewer vaginal deliveries (25.9% vs 29.6%; aRR 0.93, 95%CI 0.88-0.98, p=0.01) compared with the pre-pandemic group (Table 2).

Rates of SCN admissions were significantly lower in exposure 1 (38.5% vs 43.4%; aRR 0.91, 95%CI 0.87-0.95, p<0.001) (Table 3). However, there were no significant differences in stillbirths when births<24 weeks, TOPs and congenital anomalies were excluded (1.2% vs 0.9% aRR 1.00, 95%CI 0.99–1.01, p=0.64). Rates of FGR, NICU admissions and low Apgar scores were also unchanged between the two groups.

### Exposure 2 group versus pre-pandemic group

#### Primary outcomes

There were no significant differences in the rates of total PTB<37 weeks or all-cause stillbirths in the exposure 2 group compared with the pre-pandemic group (Tables 2 and 3, respectively).

#### Secondary outcomes

There was no significant change in rates of PTB<34 weeks or iatrogenic births for suspected fetal compromise in exposure 2 compared with the pre-pandemic group (Table 2). Adjusted stillbirths were also unchanged (0.8% vs 0.9%, aRR 0.99, 95%CI 0.99-1.00, p=0.15) (Supplementary Table 2).

However, exposure 2 was associated with significantly higher rates of PTB<28 weeks (7.2% vs 4.8%; aRR 1.03, 95%CI 1.01-1.05, p=0.04) with higher rates of both spontaneous and iatrogenic PTBs (Table 2). The GA distributions of the pre-pandemic, exposure 1 and exposure 2 cohorts, per pregnancy, are displayed in Figure 3, demonstrating the shift to more term births in exposure 1 and more extreme PTBs in exposure 2.

There were higher NICU admissions in exposure 2 compared with the pre-pandemic period (25.0% vs 19.6%; aRR 1.06, 95%CI 1.03-1.10, p<0.001). There were fewer SCN admissions (35.0% vs 43.4%: aRR 0.87, 95%CI 0.84-0.91, p<0.001) and fewer infants born with congenital defects (4.0% vs 4.4%; aRR 0.97, 95%CI 0.96-0.99, p<0.001 (Table 3).

#### Subgroup Analysis by fetal sex concordance

There were significantly fewer PTBs<34 weeks among sex-discordant twins compared with sex-concordant twins (17.7% vs 25.5%, p<0.001) (combined pre-pandemic and exposure cohorts). Similarly, iatrogenic births for suspected fetal compromise were lower for sex-discordant than sex-concordant twins (15.8% vs 19.4%, p<0.001) (Table 4). This was an expected finding that is likely explained by the recommended practice of delivering MCDA twins >37 weeks.

#### Sensitivity analyses

There were 2069 women birthing twins when we excluded births<24 weeks, TOPs, and infants with congenital anomalies (8.7% of births excluded). The primary and secondary outcomes for exposure 1 and exposure 2 trended in the same directions (Supplementary Tables 1 and 2). There were fewer PTB<37 weeks in the exposure 1 group (62.6% vs 67.1%, aRR 0.94 95%CI 0.88-0.99, p=0.04) compared to the pre-pandemic group (Supplementary Table 1).

The exposure 2 group showed a trend to higher total PTB 37 weeks compared with the pre-pandemic group (70.3% vs 67.1%; aRR 1.05 95%CI 0.98-1.10, p=0.09) (Supplementary Table 1). There were also significantly higher rates of total PTB<34 weeks (24.3% vs 20.0%; aRR 1.06, 95%CI 1.01-1.11, p=0.01) and iatrogenic PTB<34 weeks (13.7% vs 10.8%; aRR 1.04, 95%CI 1.01-1.07, p=0.04) (Supplementary Table 1).

In the second sensitivity analysis excluding sex-discordant twins, we included 1579 pregnancies (24.2% excluded). Total PTB<37 weeks were significantly lower in exposure 1 group compared to the pre-pandemic group (68.5% vs 73.2%; aRR 0.93, 95%CI 0.87–0.99, p=0.03) (Supplementary Table 3). There was no significant difference in the rates of stillbirth (Supplementary Table 4).

## COMMENT

### Principal findings

Our study provides a detailed analysis of twin outcomes during the pandemic over two years of restrictions in a high-income setting. During the first exposure period with the strictest restrictions, we found significantly lower rates of PTB<37 and PTB<34 weeks compared with the pre-pandemic period. This was accompanied by a trend to lower iatrogenic PTB, but reassuringly, no significant differences in stillbirths.

The lower PTB rate during exposure 1 was not observed in exposure 2, suggesting the two stages of the pandemic had different impacts on twin perinatal outcomes. Rather, during exposure 2, there was a significant shift to more extreme PTB<28 weeks and NICU admissions.

### Results in the context of what is known

There are numerous studies on the impact of COVID-19 lockdowns on pregnancy outcomes in singleton or mixed populations, with varied results due to population, lockdown stringency and methodological heterogeneity. A recent meta-analysis concluded that the pandemic was associated with reduced PTB<37 weeks in high-income settings.^27^ Similarly, an international study of 26 countries found that the first 3 months of lockdown was associated with 3-4% relative reduction in PTB in high and upper-middle income countries. In both studies there was no associated increase in stillbirth.^28^ This was not our local experience, where we observed a reduction in iatrogenic PTB among singletons in Melbourne, in association with more preterm stillbirths.^7^

Few studies have specifically examined twin perinatal outcomes during the pandemic. A Chinese study of 210676 multiple births demonstrated an immediate 2.8% absolute reduction in PTB (95%CI_1.09%-4.51%, p=0.002) in the first month of restrictions without an increase in stillbirths. The reduction was associated with reduced spontaneous, but not iatrogenic PTB and was predominantly driven by late PTB. The changes did not persist two months after the implementation of restrictions.^29^ Our study similarly demonstrates a lower PTB rate during early restrictions (exposure 1), with a return to baseline rates by the second year of the pandemic (exposure 2). In contrast, a Danish quasi-experimental study found no difference in twin PTB rates during the COVID-19 lockdown and mitigation periods compared to the pre-pandemic period.^30^ However, unlike this Danish study we used cLMP to define our cohorts and had a longer study period with much larger sample size.

Due to the higher perinatal morbidity and mortality rate for twin pregnancies, our health care sector was concerned that pandemic disruptions to maternity care may have hindered early diagnosis and timely intervention for twin-related complications, such as TTTS. International investigators have reported adverse impacts on fetal surveillance and timely management of TTTS leading to poorer survival.^31^ However, an international survey of 561 women with MCDA pregnancies, including Australian women, showed most women’s care adhered to established guidelines during the pandemic. Some respondents felt their care was *better*, as high-risk pregnancies were prioritized during a time of resource rationing, corroborated by 4.7% more double MCDA twin survivors compared to the pre-pandemic period (p=0.04).^32^ Although we were unable to analyze our results by chorionicity, our subgroup analysis supports these survey results as we did not detect higher rates of all-cause stillbirths among sex-concordant twins compared with sex-discordant twins. However, the lower number of twin pregnancies in exposure 1 may reflect reduced access to fertility services, due to the Victorian Government implementing restrictions on elective surgery including in-vitro fertilization from March 2020 with 85% of pre-COVID activity resumed in September 2020.^33, 34^

### Clinical Implications

A state-wide study of twins in our population conducted before the pandemic demonstrated a steady increase in PTBs, largely attributable to increases in late and iatrogenic PTB.^35^ It is well-established that late PTBs are associated with higher neonatal morbidity and mortality compared to term infants^36^ as well as substantial long-term health and educational costs.^37^ Our results by sex-concordance show that our local practice reflects current recommendations of planned birth at 37^+0^ weeks for uncomplicated dichorionic pregnancies and at 36^+0^ weeks for uncomplicated MCDA pregnancies^8^ with the median GA of 36^+0^ weeks for sex-concordant twins and 36^+6^ weeks for sex-discordant twins. That iatrogenic PTB for suspected fetal compromise was significantly lower during exposure 1 without any significant differences in stillbirths or adverse neonatal outcomes (including FGR) suggests a review of current guidelines may be warranted as there may be scope to safely reduce iatrogenic PTB in uncomplicated twin pregnancies.

The 25.0% NICU admission rate during exposure 2 translates to 66 more infant admissions than expected. Exposure 2 was marked by increased mobility compared with 2020, including for non-essential shopping, recreation, and workplace attendance.^38^ Reasons underlying the higher rates of spontaneous PTB<28 weeks and NICU admissions in exposure 2 are unclear and ongoing monitoring of extreme PTB is warranted.

### Research implications

The rise in twin PTB in Australia, with no associated improvements in perinatal mortality, raises concerns that we are not achieving the optimal balance between avoiding stillbirth and minimizing the consequences of iatrogenic PTB.^39, 40^ Further research is needed on the optimum time for delivery of uncomplicated twin pregnancies.

### Strengths and Limitations

Melbourne offered a unique amalgamation of stringent and prolonged restrictions, low COVID-19-related obstetric morbidity and mortality, and a well-resourced healthcare system, making it the ideal setting to study the impact of lockdown on perinatal outcomes. The size and definition of our cohort, detailed patient-level information, and complete capture of all public hospital births overcomes many limitations in single-center and population-based datasets.

We were unable to capture home and private hospital births. Private hospital births differ in outcomes including higher rates of CS, undetected FGR and IOL.^40^ However, any high-risk pregnancy with expected PTB<31^+0^ weeks or with severe fetal or maternal complications would be transferred to one of the public tertiary hospitals included in this study. Additionally, pregnancies ending<20^+0^ weeks were not routinely recorded in maternity data, so we could not detect the effect on lockdown on these pregnancies.^17^ Finally, we used sex-concordance as an imperfect surrogate for chorionicity as not all hospitals reported chorionicity. Our results did not indicate a rise in perinatal complications in this group, but we may have had insufficient power to detect a difference in MCDA outcomes by using a mixed population of sex-concordant twins.

We treated each infant in a twin pair as an independent data point, not accounting for the non-independence of infants from twin pregnancies for outcomes such as stillbirth. The effect of fetal demise on the co-twin is expected to be greater in monochorionic than dichorionic pregnancies, as the risk of fetal death in the co-twin is 12% compared to 4%, respectively.^42^ Several methods have been described to account for the non-independence of infants from multiple pregnancies^43^ however, we chose to assume independence which has the advantage of maintaining the size and power of our dataset. While our estimates of risk ratio are unaffected by this assumption, our confidence intervals for infant outcomes such as stillbirths and FGR may be narrower than the true intervals.^43^

### Conclusion

Melbourne’s first year of lockdown was associated with fewer premature twin births, including both iatrogenic and spontaneous PTB. This decline was not associated with significant differences in adverse perinatal outcomes, such as stillbirth or FGR. These lower rates of PTB were not sustained into the second lockdown period. Our findings provide important insights into the influences on PTB and suggest scope to review our approach to the optimal timing of delivery and provision of antenatal care for twin pregnancies.

## CONTRIBUTORS

LH, SPW, JMS, CLW, DLR, BWM, KRP and SP conceived of the original collaboration, LH designed, coordinated, and acquired funding for the CoMaND collaboration; LH and MM drafted the data analysis plan, and LH, SPW, BWM, JF, PMS, CLW, KRP, JMS, NP, DLR, SP, MBM, JMM edited and/or approved the analysis plan; LH, SPW, JMS, DLR, CLW, PMS, JF, MBM collected the primary data; MBM and JMM performed the data cleaning and coding; LH, MBM and JMM performed the data analysis; JMM, MBM and LH created the tables and figures; JMM, LH and MBM performed the literature review; JMM and LH wrote the primary manuscript; all authors reviewed and edited the draft manuscript for scientific content and approved the final submitted manuscript.

## DATA SHARING

Non-identifiable individual participant data is available to researchers affiliated with a recognizedz academic institution following requests to the Austin Health Human Research Ethics Committee and the Mercy Health Human Research Ethics Committee. The study investigators may contribute aggregate and non-identifiable individual patient data to national and international collaborations whose proposed use of the data has been ethically reviewed and approved by an independent committee and following signing of an appropriate research collaboration agreement with the University of Melbourne.

## Data Availability

All data produced in the present study are available upon reasonable request to the authors

## ACKNOWLEDGEMENTS

The health services and individual hospitals contributing to the Collaborative Maternity and Newborn Dashboard for the COVID-19 Pandemic are:

· Mercy Health (Mercy Hospital for Women, Werribee Mercy Hospital)

· The Royal Women’s Hospital, The Women’s at Sandringham

· Monash Health (Monash Medical Centre, Casey Hospital, Dandenong Hospital)

· Northern Health (The Northern Hospital)

· Western Health (Joan Kirner Women’s and Children’s Hospital)

· Eastern Health (Box Hill Hospital, The Angliss Hospital)

· Peninsula Health (Frankston Hospital)

We thank the health service data managers and research midwives (Tania Fletcher, Lynn Rigg, Michelle Knight, Eleanor Johnson, Abby Monaghan, Pauline Hamilton, Roshanee Perera) for their assistance with primary data collection and Dr Andrew Goldsack for his assistance with coding of the indications for iatrogenic births.

