## Supplemental tables 1-4 for "Reduction in Spontaneous and Iatrogenic Preterm Births in Twin Pregnancies During COVID-19 Lockdown in Melbourne, Australia: A Multicenter Cohort Study"

**APPENDIX**

**Supplementary Table 1**: Primary and secondary outcomes among all pregnancies in the pre-pandemic, exposure 1 and exposure 2 groups excluding pregnancies with births<24 weeks, terminations of pregnancy and infants with congenital anomalies.

| Outcomes | Pre-pandemic | | Exposure 1 | | Exposure 2 | | Exposure 1 aRR | | | Exposure 2 aRR | | |
|---|---|---|---|---|---|---|---|---|---|---|---|---|
| | **n=1120** | **54%** | **n=393** | **19%** | **n=556** | **27%** | **aRR** | **95% CI** | **P value** | **aRR** | **95% CI** | **P value** |
| **Preterm birth <37 wk** | | | | | | | | | | | | |
| Total | 751 | 67.1 | 246 | 62.6 | 391 | 70.3 | 0.94 | 0.88-0.99 | 0.04 | 1.05 | 0.98-1.10 | 0.09 |
| Spontaneous | 215 | 19.2 | 74 | 18.8 | 117 | 21.0 | 0.96 | 0.92-1.01 | 0.15 | 1.00 | 0.96-1.05 | 0.98 |
| Iatrogenic | 536 | 47.9 | 172 | 43.8 | 274 | 49.3 | 0.97 | 0.91-1.04 | 0.43 | 1.05 | 0.99-1.11 | 0.12 |
| **Preterm birth <34 wk** | | | | | | | | | | | | |
| Total | 224 | 20.0 | 71 | 18.1 | 135 | 24.3 | 0.96 | 0.91-1.01 | 0.14 | 1.06 | 1.01-1.11 | 0.01 |
| Spontaneous | 103 | 9.2 | 37 | 9.4 | 59 | 10.6 | 0.99 | 0.95-1.02 | 0.48 | 1.02 | 0.99-1.05 | 0.22 |
| Iatrogenic | 121 | 10.8 | 34 | 8.7 | 76 | 13.7 | 0.98 | 0.94-1.02 | 0.24 | 1.04 | 1.01-1.07 | 0.04 |

| | | | | | | | | | | | | |
|---|---|---|---|---|---|---|---|---|---|---|---|---|
| **Preterm birth <28 wk** | | | | | | | | | | | | |
| Total | 29 | 2.6 | 13 | 3.3 | 22 | 4.0 | 1.00 | 0.97-1.03 | 0.96 | 1.01 | 0.99-1.04 | 0.21 |
| Spontaneous | 16 | 1.4 | 6 | 1.5 | 11 | 2.0 | 1.00 | 0.98-1.01 | 0.91 | 1.01 | 1.00-1.03 | 0.07 |
| Iatrogenic | 13 | 1.2 | 7 | 1.8 | 11 | 2.0 | 1.00 | 0.98-1.01 | 0.60 | 1.01 | 0.99-1.02 | 0.31 |
| **Iatrogenic birth for suspected fetal compromise** | | | | | | | | | | | | |
| Total | 230 | 20.5 | 49 | 12.5 | 110 | 19.8 | 0.93 | 0.88-0.98 | 0.00 | 0.99 | 0.95-1.04 | 0.77 |
| ≥37 wk | 50 | 13.6 | 9 | 6.1 | 25 | 15.2 | 0.93 | 0.87-1.00 | 0.05 | 1.02 | 0.95-1.09 | 0.20 |
| <37 wk | 180 | 24.0 | 40 | 16.3 | 85 | 21.7 | 0.93 | 0.87-1.00 | 0.04 | 0.98 | 0.93-1.04 | 0.71 |
| **Mode of birth** | | | | | | | | | | | | |
| Vaginal | 327 | 29.2 | 100 | 25.5 | 158 | 28.4 | 0.93 | 0.88-0.98 | 0.01 | 0.96 | 0.91-1.01 | 0.09 |
| Caesarean section after labor onset | 378 | 33.8 | 131 | 33.3 | 208 | 37.4 | 0.97 | 0.91-1.03 | 0.28 | 1.02 | 0.97-1.07 | 0.49 |

| | | | | | | | | | | | | |
|---|---|---|---|---|---|---|---|---|---|---|---|---|
| Caesarean section with no labor | 415 | 37.1 | 162 | 41.2 | 190 | 34.2 | 1.11 | 1.05-1.18 | <0.001 | 1.02 | 0.97-1.08 | 0.38 |
| **Pregnancy care indicators** | | | | | | | | | | | | |
| First antenatal visit ≤12 wk | 668 | 59.6 | 268 | 68.2 | 389 | 70.0 | 1.04 | 0.98-1.11 | 0.18 | 1.05 | 0.99-1.10 | 0.23 |
| Severe PPH > 1000ml | 124 | 11.1 | 36 | 9.2 | 57 | 10.3 | 0.95 | 0.91-0.99 | 0.01 | 0.97 | 0.94-1.00 | 0.62 |

aRR, adjusted odds ratio; CI, confidence interval; PPH, post-partum haemorrhage; wk, weeks.

**Supplementary Table 2:** Infant outcomes in the pre-pandemic, exposure 1 and exposure 2 groups excluding pregnancies with congenital anomalies, terminations of pregnancy and births <24 weeks

| Outcomes | Pre-pandemic | | Exposure 1 | | Exposure 2 | | Exposure 1 aRR | | | Exposure 2 aRR | | |
|---|---|---|---|---|---|---|---|---|---|---|---|---|
| | **n=2438** | **58%** | **n=866** | **19%** | **n=1230** | **27%** | **aRR** | **95%CI** | **P value** | **aRR** | **95%CI** | **P value** |
| Adjusted stillbirths | 21 | 0.9 | 9 | 1.2 | 9 | 0.8 | 1.00 | 0.99-1.01 | 0.64 | 0.99 | 0.99-1.00 | 0.15 |
| Fetal growth restriction | 133 | 5.9 | 43 | 5.5 | 59 | 5.3 | 1.00 | 0.98-1.02 | 0.86 | 0.99 | 0.98-1.01 | 0.58 |
| SCN admissions | 998 | 44.6 | 316 | 40.2 | 415 | 37.3 | 0.91 | 0.87-0.95 | <0.001 | 0.87 | 0.84-0.91 | 0.00 |
| NICU admissions | 407 | 18.2 | 148 | 18.8 | 259 | 23.3 | 1.00 | 0.96-1.04 | 0.99 | 1.08 | 1.04-1.11 | 0.00 |
| 5-minute APGAR <7 (term infants) | 26 | 3.5 | 14 | 4.8 | 15 | 4.6 | 0.99 | 0.97-1.02 | 0.72 | 1.00 | 0.97-1.03 | 0.89 |

*aRR*, adjusted risk ratio; *CI*, confidence interval; *SCN*, special care nursery; *NICU*, neonatal intensive care unit.

**Supplementary Table 3**:  Primary and secondary outcomes for sex-concordant twin pregnancies in the pre-pandemic, exposure 1 and exposure 2 groups

| Outcomes | Pre-pandemic | | Exposure 1 | | Exposure 2 | | Exposure 1 aRR | | | Exposure 2 aRR | | |
|---|---|---|---|---|---|---|---|---|---|---|---|---|
| | **n=829** | **54%** | **n=302** | **19%** | **n=448** | **27%** | **aRR** | **95%CI** | **P value** | **aRR** | **95%CI** | **P value** |
| **Preterm birth <37 wk** | | | | | | | | | | | | |
| Total | 607 | 73.2 | 207 | 68.5 | 340 | 75.9 | 0.93 | 0.87- 0.99 | 0.03 | 1.03 | 0.97-1.09 | 0.32 |
| Spontaneous | 166 | 20.0 | 57 | 18.9 | 92 | 20.5 | 0.95 | 0.89-1.01 | 0.07 | 0.98 | 0.93-1.03 | 0.41 |
| Iatrogenic | 441 | 53.2 | 150 | 49.7 | 248 | 55.4 | 0.98 | 0.91-1.06 | 0.64 | 1.05 | 0.99-1.12 | 0.13 |
| **Pre-term birth <34 wk** | | | | | | | | | | | | |
| Total | 205 | 24.7 | 68 | 22.5 | 133 | 29.7 | 0.95 | 0.89-1.01 | 0.08 | 1.04 | 0.99-1.10 | 0.13 |
| Spontaneous | 86 | 10.4 | 34 | 11.3 | 49 | 10.9 | 0.99 | 0.94-1.03 | 0.58 | 1.00 | 0.96-1.04 | 0.85 |
| Iatrogenic | 119 | 14.4 | 34 | 11.3 | 84 | 18.8 | 0.96 | 0.91-1.01 | 0.10 | 1.04 | 1.00-1.09 | 0.07 |

| | | | | | | | | | | | | |
|---|---|---|---|---|---|---|---|---|---|---|---|---|
| **Pre-term birth <28 wk** | | | | | | | | | | | | |
| Total | 39 | 4.7 | 17 | 5.6 | 34 | 7.6 | 0.98 | 0.95-1.01 | 0.16 | 1.01 | 0.98-1.04 | 0.40 |
| Spontaneous | 16 | 1.9 | 8 | 2.7 | 16 | 3.6 | 1.00 | 0.98-1.02 | 0.83 | 1.02 | 1.00-1.03 | 0.11 |
| Iatrogenic | 23 | 2.8 | 9 | 3.0 | 18 | 4.0 | 0.98 | 0.95-1.00 | 0.08 | 1.00 | 0.97-1.02 | 0.72 |
| **Iatrogenic birth for fetal compromise** | | | | | | | | | | | | |
| Total | 175 | 21.1 | 44 | 14.6 | 98 | 21.9 | 0.94 | 0.89- 1.00 | 0.04 | 1.01 | 0.96-1.06 | 0.77 |
| ≥37 Weeks | 27 | 12.2 | 6 | 6.3 | 18 | 16.7 | 0.96 | 0.89-1.05 | 0.41 | 1.05 | 0.97-0.97 | 0.20 |
| <37 Weeks | 148 | 24.4 | 38 | 18.4 | 80 | 23.5 | 0.94 | 0.87-1.01 | 0.09 | 0.99 | 0.93-1.05 | 0.71 |
| **Mode of birth** | | | | | | | | | | | | |
| Vaginal | 249 | 30.0 | 78 | 25.8 | 127 | 28.4 | 0.92 | 0.87-0.99 | 0.02 | 0.96 | 0.91-1.01 | 0.13 |
| Cesarean section after labor onset | 287 | 34.6 | 99 | 32.8 | 176 | 39.3 | 0.96 | 0.89-1.03 | 0.23 | 1.02 | 0.96-1.08 | 0.58 |

| | | | | | | | | | | | | |
|---|---|---|---|---|---|---|---|---|---|---|---|---|
| Ceasarean section with no labor | 293 | 35.3 | 125 | 41.4 | 145 | 32.4 | 1.13 | 1.05-1.21 | <0.001 | 1.03 | 0.97-1.09 | 0.41 |
| **Pregnancy Care Indicators** | | | | | | | | | | | | |
| First antenatal visit ≤12 weeks | 491 | 59.2 | 204 | 67.6 | 311 | 69.4 | 1.02 | 0.95-1.09 | 0.62 | 1.04 | 0.98-1.10 | 0.23 |
| Severe PPH > 1 L | 81 | 9.8 | 25 | 8.3 | 49 | 10.9 | 0.95 | 0.90-0.99 | 0.01 | 0.99 | 0.95-1.03 | 0.62 |

*aRR*, adjusted odds ratio; *CI*, confidence interval; *PPH*, post-partum haemorrhage; *wk*, weeks.

**Supplementary Table 4**: Infant outcomes among sex-concordant pregnancies in the pre-pandemic, exposure 1 and exposure 2 groups.

| Outcomes | Pre-pandemic | | Exposure 1 | | Exposure 2 | | Exposure 1 aRR | | | Exposure 2 aRR | | |
|---|---|---|---|---|---|---|---|---|---|---|---|---|
| | n=1658 | 54% | n=604 | 19% | n=896 | 27% | aRR | 95%CI | P value | aRR | 95%CI | P value |
| All-cause stillbirth | 25 | 1.5 | 11 | 1.8 | 24 | 2.7 | 1.00 | 0.99-1.02 | 0.48 | 1.00 | 0.99-1.01 | 0.72 |
| Fetal growth restriction | 103 | 6.2 | 32 | 5.3 | 53 | 5.9 | 0.99 | 0.97-1.02 | 0.64 | 1.00 | 0.98-1.02 | 0.99 |
| SCN Admission | 730 | 44.1 | 236 | 39.1 | 308 | 34.4 | 0.95 | 0.91-1.00 | 0.04 | 0.91 | 0.88-0.95 | <0.001 |
| NICU Admission | 357 | 21.5 | 139 | 23.0 | 238 | 26.6 | 1.01 | 0.97-1.06 | 0.62 | 1.07 | 1.03-1.11 | <0.001 |
| 5-minute Apgar <7 at term | 16 | 3.6 | 12 | 6.3 | 9 | 4.2 | 1.01 | 0.97-1.05 | 0.64 | 1.00 | 0.97-1.04 | 0.80 |

*aRR*, adjusted risk ratio; *CI*, confidence interval; *SCN*, special care nursery; *NICU*, neonatal intensive care unit.
